# REM sleep EEG slowing reflects brain cholinergic denervation in aging and Mild Cognitive Impairment

**DOI:** 10.1101/2025.04.28.25326545

**Authors:** Claire André, Marc-André Bédard, Véronique Daneault, Rebekah Wickens, Jean-Paul Soucy, Dominique Lorrain, Célyne Bastien, Carol Hudon, Nicola Andrea Marchi, Jacques Montplaisir, Nadia Gosselin, Julie Carrier

## Abstract

Brain cholinergic denervation is among the earliest manifestations of Alzheimer’s disease, and rapid eye movement (REM) sleep alterations have also been described early in the course of the disease. While cholinergic activity supports cortical activation during REM sleep, direct evidence for a link between cholinergic degeneration and early REM sleep alterations in humans is still lacking. Here, we tested the long-standing hypothesis that early cholinergic denervation may be associated with REM sleep EEG slowing in older adults with and without Mild Cognitive Impairment.

Twenty-four older participants (mean age: 71.29 ± 4.85 years; 58.33% women; 25% participants with amnestic Mild Cognitive Impairment) without dementia or moderate-to-severe obstructive sleep apnea underwent a night of in-laboratory polysomnography, comprehensive neuropsychological evaluation, structural MRI and molecular PET imaging with [^18^F]-Fluoroethoxybenzovesamicol (FEOBV), known for its sensitivity to quantify brain cholinergic innervation. Voxel-wise multiple regressions assessed the associations between REM sleep characteristics (i.e., REM sleep percentage, relative theta power and EEG slowing ratios, defined as [delta + theta]/[alpha + beta] power) and FEOBV-PET standard uptake value ratio maps corrected for partial volume effects, controlling for sex. Given that FEOBV uptake was higher in women compared to men, we also performed exploratory sex-stratified analyses adjusted for age.

Higher REM sleep EEG slowing over frontal and parietal derivations was significantly associated with cortical cholinergic denervation, notably in fronto-parietal areas and the medial temporal lobe (*P<0.005* level, combined with a cluster-level family-wise error correction). Exploratory sex-stratified analyses revealed that REM sleep EEG slowing was associated with cholinergic denervation in medial temporal regions in women, and neocortical regions in men.

These findings provide the first direct *in vivo* evidence that REM sleep EEG slowing could represent a sensitive marker of cortical cholinergic denervation in older adults, prior to dementia onset. Thus, quantitative REM sleep EEG may constitute a promising marker for early diagnosis and disease-modifying interventions in Alzheimer’s disease.

## 1. Introduction

Alzheimer’s disease is characterized by the progressive accumulation of amyloid and tau pathologies in the brain, leading to neurodegeneration and beginning decades before the first clinical symptoms appear. The cholinergic system is particularly vulnerable in Alzheimer’s disease, with the loss of cholinergic terminals occurring early in the course of the disease^1–4^. Indeed, it has been shown that basal forebrain cholinergic nuclei undergo volume loss before the medial temporal lobe and neocortex^2,4^. As basal forebrain nuclei are the main source of cortical cholinergic projections^5,6^, their early degeneration is likely resulting in substantial cortical cholinergic denervation, which may be considered as a very good surrogate marker for Alzheimer’s disease detection and evolution.

Over the last few years, several Positron Emission Tomography (PET) tracers have been developed to assess the integrity of the cholinergic system *in vivo*, targeting key components such as the acetylcholinesterase, acetylcholine receptors or the vesicular acetylcholine transporter (VAChT). Among them, ^18^F-fluroethoxybenzovesamicol (FEOBV) is the tracer showing the best affinity and specificity for the VAChT, present in presynaptic cholinergic terminals, thereby reliably reflecting presynaptic cholinergic density^7–10^. FEOBV-PET was found very sensitive to detect subtle cholinergic loss resulting from experimental lesions of the basal forebrain or pedunculopontine nuclei in rodents^11,12^. In humans, recent FEOBV-PET studies have revealed significant cholinergic denervation in patients with Alzheimer’s disease^13,14^ or Mild Cognitive Impairment^15^, mainly in the cingulate and temporo-parietal cortex, as well as in the frontal areas, with those reductions being directly correlated with the severity of basal forebrain atrophy^3,13–15^.

Interestingly, cholinergic loss likely compromises the integrity of rapid eye movement (REM) sleep, and may contribute to the impairment of REM sleep in Alzheimer’s disease. In animal models of Alzheimer’s disease, REM sleep is often shortened and fragmented^16–18^. In humans, Alzheimer’s disease patients exhibit a significant reduction of REM sleep duration, prolonged REM sleep latency and pathological slowing of EEG rhythms during REM sleep compared to cognitively healthy controls^19^. Such REM sleep EEG slowing is already detectable in patients with amnestic Mild Cognitive Impairment^20,21^. Interestingly, it is known that cholinergic activity reaches its peak during REM sleep, supporting the characteristic high cortical EEG activity of this sleep stage^22–24^. As a consequence, it has long been hypothesized that cholinergic loss may underlie the alterations of REM sleep observed in Alzheimer’s disease^25^. In this respect, promising results reported by our team show that reduced REM sleep duration is associated with lower basal forebrain volume in individuals with amnestic Mild Cognitive Impairment^26^. However, no study to date has directly assessed whether the loss of cholinergic terminals assessed using molecular imaging directly contributes to REM sleep alterations in older adults. The recent development of FEOBV-PET, allowing for the specific in vivo quantification of presynaptic cholinergic terminals, now enables testing this long-standing hypothesis.

Here, we tested whether cholinergic denervation, quantified using FEOBV-PET imaging, was associated with REM sleep EEG alterations in older adults with and without amnestic Mild Cognitive Impairment. We hypothesized that REM sleep alterations typically observed in Alzheimer’s disease – especially REM sleep EEG slowing, lower REM sleep proportion and/or lower REM sleep theta power – would be associated with cholinergic loss, especially in fronto-temporal areas and the medial temporal lobe.

## 2. Materials & Methods

### 2.1. Participants

Twenty-five participants were recruited between 2022 and 2024 from local memory and sleep clinics and the community in Montreal (Canada) and its surroundings. Participants underwent a phone screening followed by an in-person interview to verify their potential eligibility, and were then invited for a comprehensive neuropsychological assessment and a full night in-laboratory polysomnography. Inclusion criteria were being 65 years old or above, fluent in French or English, having at least seven years of education and showing preserved autonomy in daily life. Exclusion criteria were the presence of dementia, the presence or history of major neurological disorders (e.g., epilepsy, traumatic head injury or encephalopathy), the presence of a psychiatric disorder diagnosed according to DSM-5 criteria (e.g., major depression or anxiety), a body mass index greater than 35 kg/m^2^, smoking, alcohol or drug abuse, heavy consumption of caffeinated beverages, uncontrolled diabetes or hypertension, presenting with sleep disorders (e.g., moderate-to-severe obstructive sleep apnea assessed with an apnea-hypopnea index ≥15, insomnia, REM sleep behaviour disorder), the presence of cerebrovascular or pulmonary diseases (e.g., history of stroke, chronic obstructive pulmonary disease), the use of medication affecting sleep, cognition, mood or brain functioning (e.g., antidepressants, hypnotics, opioids), or any reason preventing from performing an MRI or PET scan (e.g., claustrophobia, metallic implants, annual nuclear exposure over 50 mSv). The protocol was approved by the ethics committee of the CIUSSS du Nord-de-l’île-de-Montréal (MP-32-2018-1537), and a written informed consent was obtained from each participant prior to the examinations, according to the declaration of Helsinki. Participants underwent a neuropsychological evaluation, MRI scan, FEOBV-PET scan and a full-night in-laboratory polysomnography in a mean interval of 5.75 ± 4.04 months.

### 2.2. Neuropsychological testing and questionnaires

A comprehensive neuropsychological assessment was performed, encompassing global cognitive functioning (Montreal Cognitive Assessment; MoCA) and four specific cognitive domains: 1) learning and memory (Rey Auditory Verbal Learning test; Logical Memory from the Wechsler Memory Scale third edition; immediate and delayed recall of the Rey Complex Figure Test), 2) attention and executive functions (Digit Symbol Substitution Task-Coding of the WAIS-IV, Alpha-Span, Trail Making Test, Verbal fluency, Stroop Color-Word Test), 3) language (Boston Naming Test) and 4) visuo-spatial functions (Copy of the Rey Complex Figure Test, Lines orientation subtest of the Birmingham Object Recognition Battery). A clinical cognitive diagnosis was established by consensus between senior neuropsychologists, based on neuropsychological performance. As previously described^26^, a cognitive domain was considered altered when participants presented with two or more Z-scores ≤1.5 standard deviation (SD) in a given cognitive domain, or if participants presented a MoCA score <26 accompanied by one Z-score ≤1.5 SD in at least two cognitive domains including memory. Following these criteria, participants for whom all cognitive domains were objectively preserved were classified as cognitively unimpaired *(n=18)*, and those with at least one impaired cognitive domain including memory were classified as having possible amnestic Mild Cognitive Impairment *(n=6)*. No participant presented with a non-amnestic Mild Cognitive Impairment profile.

In addition to cognitive tests, participants completed several questionnaires: the Geriatric Anxiety Inventory^27^, the Geriatric Depression Scale^28^, the Activities of Daily Living Scale - Mild Cognitive Impairment version^29^, the Epworth Sleepiness Scale^30^, the Vascular Burden Index^31^ and the Cognitive Complaint Questionnaire (“Questionnaire de Plainte Cognitive”, QPC)^32^.

### 2.3. Polysomnography recording

We performed a full-night in-laboratory polysomnography using a Natus system (Brain Monitor, Trex and Embla NDx; bandpass 0.3-200 Hz, digitized at a sampling rate of 512 Hz), including 12 EEG electrodes placed on the scalp according to the international 10-20 system (F3, F4, C3, C4, T3, T4, T5, T6, P3, P4, O1, O2; referenced to A1-A2), an electrooculogram, electrocardiogram, and chin and leg electromyograms. We recorded respiration and oxygen saturation using oronasal canula and thermistors, thoraco-abdominal belts and a finger pulse oximeter. Sleep stages and respiratory events were scored in 30-second epochs by certified medical electrophysiology technologists at the Montreal site, according to standard international criteria of the American Academy of Sleep Medicine^33^. Standard sleep variables were extracted from the scoring of sleep recordings (i.e., sleep onset latency, total sleep time, sleep efficiency, wake after sleep onset, the proportion of time spent in each sleep stage, the number of awakenings per hour of sleep, and REM sleep onset latency). Regarding respiratory events, sleep apneas were defined as drops of ≥90% of airflow for a minimum of 10 seconds, and hypopneas were defined as a ≥30% reduction of airflow for a minimum of 10 seconds, followed by a cortical arousal and/or a ≥3% oxygen desaturation. Obstructive sleep apnea severity was estimated by the apnea-hypopnea index, corresponding to the number of apneas and hypopneas per hour of sleep. All participants presenting with an apnea-hypopnea index ≥15, indicative of untreated moderate-to-severe sleep apnea, were excluded. Lastly, periodic limb movement during sleep (PLMS) were also scored based on the 2012 American Academy of Sleep Medicine criteria^33^, with leg movements occurring between -3.5 to +8.0 seconds around the end of an apnea or hypopnea not being scored as PLMS^34^.

### 2.4. Quantitative EEG analyses

#### 2.4.1. Sleep EEG

All quantitative EEG analyses were performed using the Snooz toolbox (https://snooztoolbox.com). Prior to spectral analyses, artifacts were first automatically detected on NREM epochs (i.e., N2 and N3 epochs)^35^, and then visually inspected by experienced sleep technologists. For REM sleep, sections of REM sleep free of eye movements or other artifacts were manually sampled from all REM sleep periods. Of note, one subject did not have artifact-free REM sleep sections and was therefore not included in quantitative REM sleep EEG analyses. For the remaining 23 subjects, the mean duration of artifact-free REM sleep sections was 117.2 ± 12.8 seconds. Welch-like spectral power analyses were performed on 4-second epochs with a cosine tapering window (spectral resolution of 0.25 Hz). Absolute REM sleep spectral power was computed on all derivations on the following frequency bands: delta (0.5-4 Hz), theta (4-8 Hz), alpha (8-13 Hz), and beta (13-32 Hz). All absolute spectral power values were normalized to total spectral power, and log-transformed relative spectral power values were used in subsequent analyses. In addition, we computed REM sleep EEG slowing ratios using absolute power values, as follows: [(delta + theta)/(alpha + beta)]. For supplementary analyses, we also computed absolute NREM sleep spectral power on N2 and N3 epochs on the following frequency bands: delta (0.6-4 Hz), theta (4-8 Hz), alpha (8-12 Hz), sigma (12-16 Hz) and beta (16-32 Hz), for all electrodes. As no hypothesis based on lateralization was made, we averaged spectral power and EEG slowing ratios bilaterally for all analyses, on frontal (F3-F4), temporal (T3-T4) and parietal (P3-P4) derivations. The analyses were focused *a priori* on these derivations given that these regions are known to exhibit REM sleep EEG slowing^36^, atrophy and hypometabolism^37^ in the early stages of Alzheimer’s disease.

#### 2.4.2. Wakefulness EEG

For secondary analyses aiming at assessing the specificity and sensitivity of our main results, spectral power was also computed on resting-state wakefulness EEG recordings. For this purpose, a 10-minute resting state waking EEG recording was performed in the morning after the night of polysomnography, one hour after wake-up time to prevent sleep inertia. Participants were lying in bed with eyes closed, and were periodically asked to open their eyes to prevent drowsiness. We selected the maximum of artifact-free EEG sections encountered for each electrode (mean duration: 149.1 ± 80.2 seconds). We extracted spectral power from artifact-free sections and computed slowing ratios following the same procedure than for REM sleep sections described above in section 2.4.1.

### 2.5. Neuroimaging examinations

A structural MRI (3T GE Discovery MR750) and a FEOBV-PET scan (GE Discovery PET/CT 690) were completed on the same day for each participant at the PERFORM Centre of Concordia University (Montréal, Canada).

A 3D T1-weighted MRI sequence was acquired with the following parameters: TR = 7.4 ms; TE = 3.06 ms; TI= 400 ms; matrix size = 256 × 256 × 176; FOV = 25.6 cm; voxel size = 1.0 mm isotropic; flip angle = 11°.

PET scan was performed three hours following a bolus intravenous injection of ^18^F-fluoroethoxybenzovesamicol (FEOBV), with a radioactivity ranging between 160 and 300 MBq. FEOBV was synthesized in the morning at the Cyclotron facility of the McConnell Brain Imaging Centre of the Montreal Neurological Institute (Montréal, Canada) and sent within an hour to the PERFORM Centre. A short low kV CT scan for attenuation correction was first performed. PET data acquisition was done in 3D list mode for a 30 min duration. PET images were reconstructed using an OSEM (Ordered Subset Expectation Maximization) algorithm with resolution recovery, into six frames of five minutes each. Reconstructed images were corrected for attenuation, scattering, random coincidences, decay and dead time. Frame-based motion correction was also performed if needed.

Image preprocessing was conducted using SPM12 (http://fil.ion.ucl.ac.uk/spm/) in Matlab R2018b (Natick, MA). For PET images, their six frames were time-averaged to create a single image. MRI scans were then linearly spatially normalized to the MNI152 template, and segmented (i.e., image registration, tissue classification, and bias correction). The same linear transformation matrices from the MRI normalization were applied to the PET images to align them with the MRI. A Müller-Gärtner partial volume correction was applied to PET data using the PETPVE toolbox (http://www.fil.ion.ucl.ac.uk/spm/ext/#PETPVE12)^38,39^. The DARTEL algorithm^40^ was used to estimate the nonlinear deformation field, and these were applied to the PET scan, further aligning the PET images to the MNI152 template space. Standardized uptake value ratio (SUVR) maps were generated from spatially normalized PET images by using supratentorial white matter as the reference region. Resulting images were smoothed using a Gaussian kernel with a full width at half-maximum of 8 mm, and were masked to exclude non-grey matter voxels from the analyses.

### 2.6. Statistical analyses

We assessed the normality and variance homogeneity of each variable using Shapiro-Wilk and Levene’s tests respectively, and all non-normal variables were log-transformed prior to statistical analyses. In order to take into account potential confounders when exploring the associations between REM sleep and cholinergic innervation, we first verified whether FEOBV uptake was affected by demographics. Therefore, we performed (i) two-sample comparisons of FEOBV SUVR maps between men and women, as well as between cognitively unimpaired versus Mild Cognitive Impairment participants, and (ii) voxel-wise simple regressions between FEOBV SUVR maps and age and education, separately. We also assessed whether REM sleep outcomes (i.e., REM sleep percentage, theta power and EEG slowing ratio) were influenced by demographic variables (i.e., age, sex, education and cognitive status), by conducting simple regression analyses, and independent two-sample T-tests for categorical variables (sex and cognitive status), combined with an FDR correction for multiple comparisons for continuous variables (age and education).

The main analyses investigated the associations between cholinergic denervation and REM sleep outcomes using whole-brain voxel-wise multiple regressions between FEOBV SUVR maps and REM sleep outcomes. For reasons of parsimony, these analyses were only controlled for sex, as sex had a significantly impact on FEOBV uptake (see **Fig. 1** and Results section 3.2). However, all results were then confirmed in a fully adjusted model, controlled for age, sex and cognitive status, based on previous literature and as age was marginally associated with REM sleep percentage only (see Results section 3.2).

**Figure 1.**
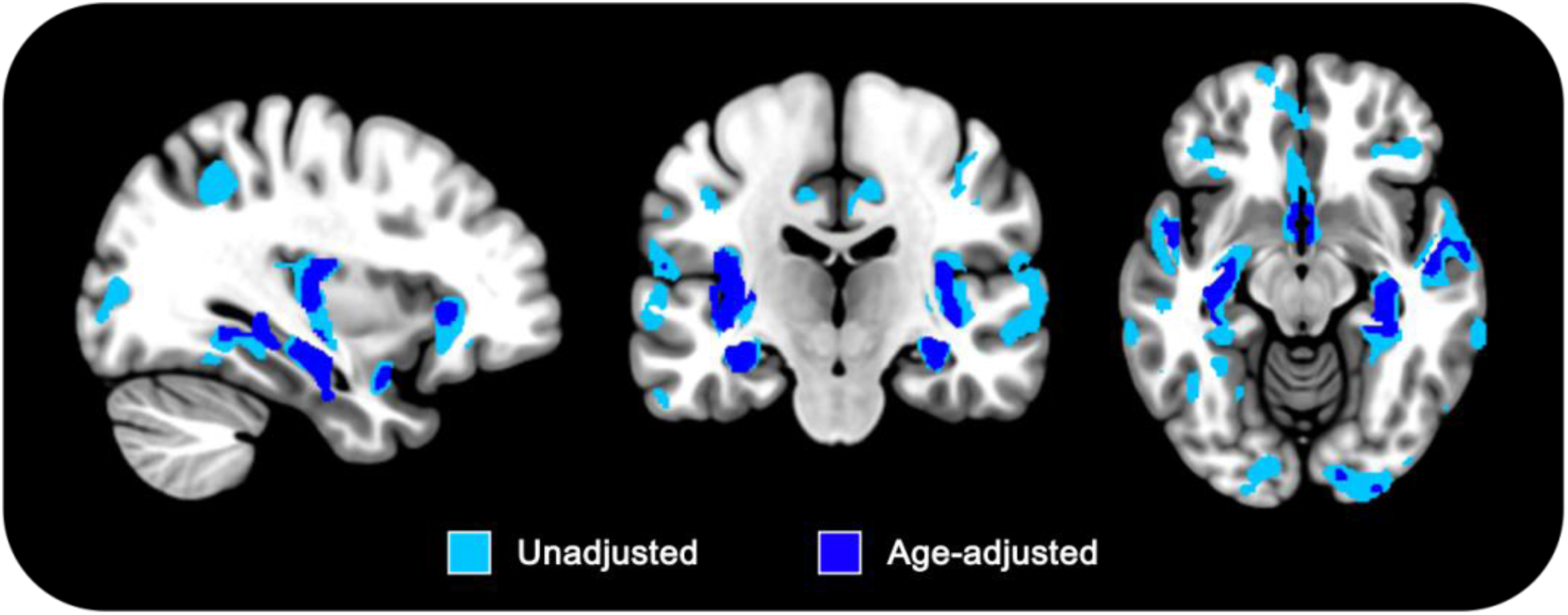
Sex differences in FEOBV uptake. Result of unadjusted (light blue) and age-adjusted (dark blue) two-sample comparisons of FEOBV uptake between women *(n=14)* and men *(n=10)* (Women>Men contrast). Significant clusters depict higher FEOBV uptake in women compared to men at the *P<0.005* uncorrected threshold, combined with a minimum cluster size of *k=100*. Detailed clusters statistics and peak coordinates are displayed in Supplementary Table 1.

As an exploratory supplementary analysis, we assessed whether our main results were replicated in sex-stratified samples, adjusting for age. Specificity analyses tested the associations with NREM sleep or wakefulness slowing ratios as independent variables.

All voxel-wise neuroimaging analyses were carried out using Matlab R2022b and SPM12 (version 7771). The main analyses were considered significant at the *P<0.005* (uncorrected) threshold, combined with a cluster-level threshold of *P<0.05* corrected for family-wise error (FWE). More exploratory analyses in subsamples (i.e., sex-stratified analyses) are presented at a more permissive threshold of *P<0.005* (uncorrected), combined with a minimum cluster size of *k=100* voxels.

## 3. Results

### 3.1. Participants

From the 25 subjects included in the present study, one participant did not complete the MRI scan due to a claustrophobia attack, and was excluded from the analytic sample. The final sample was composed of 24 subjects, including 14 women and 10 men, 18 cognitively unimpaired participants and six patients with a possible amnestic Mild Cognitive Impairment, with a mean age of 71.29 ± 4.85 years (range: 65-80 years) and a mean level of education of 16.31 ± 3.51 years (range: 10-23 years). Detailed demographic, clinical and sleep characteristics of the sample are presented in **Table 1**.

**Table 1.** Participants’ characteristics. Demographic, clinical and sleep data of the 24 participants of the study. Abbreviations: %, percentage; aMCI, amnestic Mild cognitive Impairment; NREM, non-rapid eye movement; PLMS, periodic limb movements during sleep; REM, rapid eye movement.

| Variable | Mean | Standard deviation | Minimum | Maximum |
| --- | --- | --- | --- | --- |
| Age: years | 71.29 | 4.85 | 65 | 80 |
| Sex: number (%) women |  | 14 (58.33 %) |  |  |
| Education: years | 16.33 | 3.51 | 10 | 23 |
| Body mass index: kg/m<sup>2</sup> | 25.26 | 4.33 | 18.25 | 33.25 |
| Montreal Cognitive Assessment: total score | 26.75 | 2.66 | 20 | 30 |
| Possible aMCI: number (%) |  | 6 (25 %) |  |  |
| Vascular Burden Index: total score | 0.54 | 0.88 | 0 | 3 |
| Cognitive Complaint Questionnaire: total score | 2.38 | 2.26 | 0 | 8 |
| Activities of Daily Living questionnaire: total score | 45.96 | 4.94 | 26 | 50 |
| Geriatric Depression Scale: total score | 2.25 | 2.33 | 0 | 8 |
| Geriatric Anxiety Inventory: total score | 3.83 | 4.82 | 0 | 16 |
| Epworth Sleepiness Scale: total score | 4.21 | 3.67 | 0 | 16 |
| Sleep onset latency: minutes | 23.33 | 28.43 | 0.5 | 112.5 |
| Total sleep time: minutes | 344.54 | 57.44 | 231 | 432 |
| Wake after sleep onset: minutes | 100.79 | 42.45 | 38.5 | 186 |
| Night-time awakenings: number | 32.29 | 13.70 | 10 | 61 |
| Sleep efficiency: % | 76.51 | 9.98 | 56.1 | 91.8 |
| NREM sleep stage 1: % | 15.99 | 7.00 | 4.6 | 33.8 |
| NREM sleep stage 2: % | 58.6 | 9.95 | 33.8 | 75.5 |
| NREM sleep stage 3: % | 9.175 | 9.52 | 0 | 27 |
| REM sleep: % | 16.25 | 7.04 | 0.9 | 28.3 |
| Apnea-hypopnea index: number/hour | 7.03 | 3.48 | 2.6 | 14.3 |
| Total sleep time with oxygen saturation <90%: minutes | 1.84 | 2.99 | 0 | 10.8 |
| PLMS index: number/hour | 25.84 | 26.89 | 1.04 | 76.01 |

### 3.2. Impact of demographics on REM sleep and FEOBV uptake

Women presented higher FEOBV uptake compared to men, mainly in the medial temporal lobe (e.g., hippocampus, parahippocampal gyrus, amygdala), basal forebrain, insula and temporal cortex **(Fig. 1 and Supplementary Table 1)**. FEOBV uptake did not significantly differ between cognitively unimpaired participants and those with possible amnestic Mild Cognitive Impairment, and was not significantly associated with age or education levels. The main analyses were thus controlled for sex, and exploratory sex-stratified analyses were conducted. Only a marginal negative association was observed between lower REM sleep percentage and age, which did not survive a FDR correction for multiple comparisons *(r=-0.42, p_unc._=0.043, Benjamini-Hochberg adjusted p=0.18)* **(Supplementary Fig. 1)**. There was no other significant association between REM sleep outcomes and age or education, and no significant REM sleep differences between groups stratified by sex or cognitive status **(Supplementary Fig. 1)**.

### 3.3. Associations between FEOBV uptake and REM sleep characteristics

#### 3.3.1. Whole sample

We first investigated the associations between FEOBV uptake and REM sleep characteristics using voxel-wise multiple regression analyses, controlling for sex. Higher REM sleep EEG slowing ratio over frontal and parietal derivations was significantly associated with a widespread reduction of FEOBV uptake, mainly in frontal, parietal (e.g., precuneus) and temporal areas, including the medial temporal lobe **(Fig. 2 and Supplementary Table 2)**. These associations were still significant when adding age and cognitive status as covariates **(Supplementary Fig. 2).** No significant associations were found between FEOBV uptake and REM sleep proportion or relative theta power over frontal, temporal and parietal derivations.

**Figure 2.**
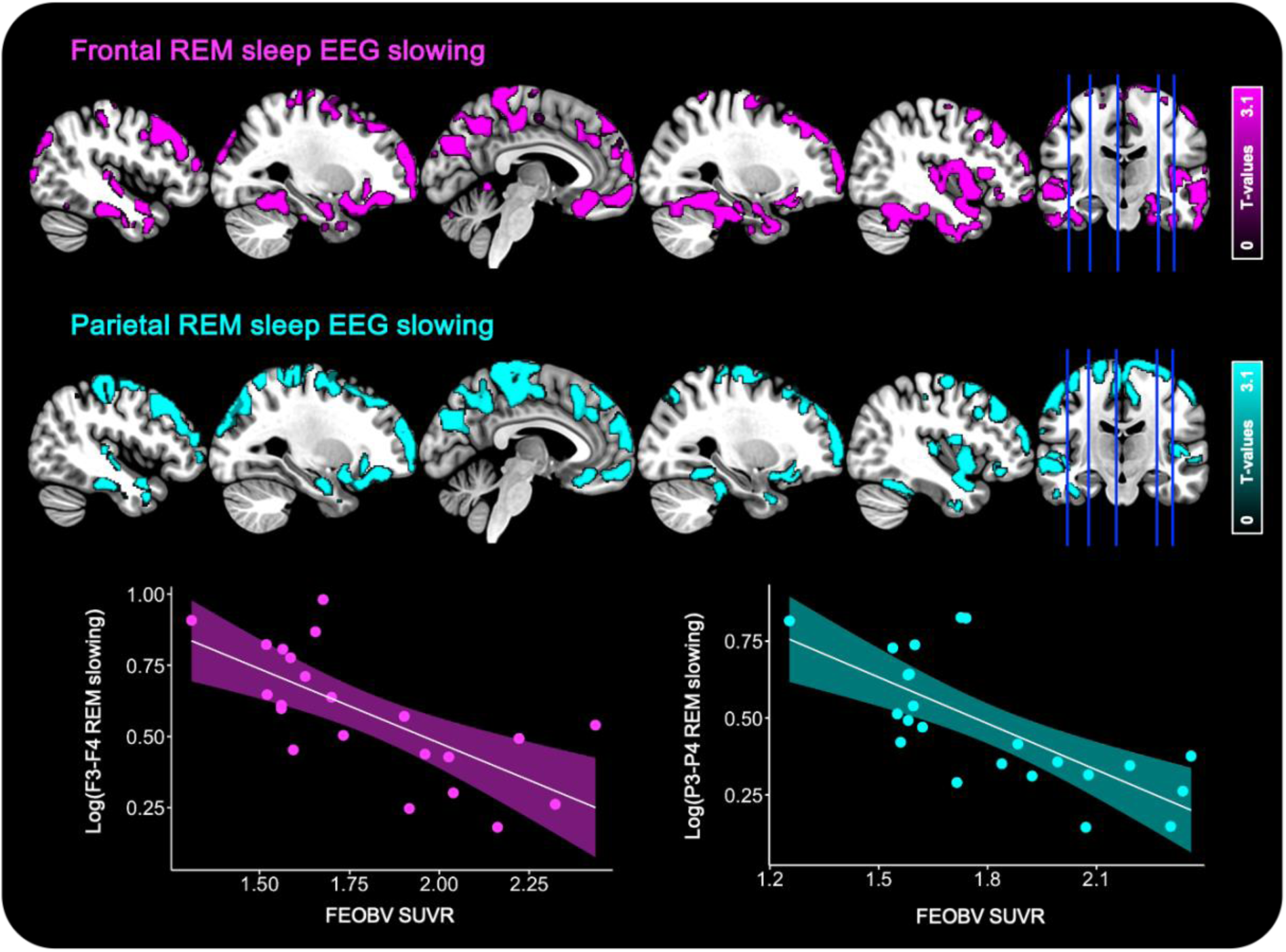
Associations between REM sleep EEG slowing ratios and FEOBV uptake. Negative voxel-wise regressions were performed between REM sleep EEG slowing ratio on F3-F4 (pink, *n=22*) and P3-P4 (cyan, *n=23*) derivations and FEOBV SUVR maps corrected for partial volume effects, controlling for sex. Results are presented at the *P<0.005* uncorrected level, combined with a cluster-level FWE correction. Detailed clusters statistics and peak coordinates are available in Supplementary Table 2.

In sensitivity and specificity analyses, we first verified whether cholinergic denervation was also associated with frontal and parietal NREM sleep or resting-state wakefulness EEG slowing ratios, controlling for sex. NREM sleep EEG slowing ratios and the parietal wake EEG slowing ratio were not associated with FEOBV uptake at the permissive *P<0.005* uncorrected threshold, combined with a minimum cluster size of *k=100*. We found a marginal negative association between the frontal wake EEG slowing ratio and FEOBV uptake in the right superior temporal pole and left superior frontal gyrus, which did not survive a FWE cluster-level correction **(Supplementary Table 3)**.

#### 3.3.2. Sex-stratified analyses

As a significant sex-difference was observed in FEOBV uptake **(Fig. 1)**, we verified whether the associations between FEOBV uptake and frontal and parietal REM sleep EEG slowing ratios were replicated in sex-stratified subgroups. As women were slightly older than men *(t=2.096, P=0.048)*, age was added as a covariate in these analyses. In women, we observed that higher REM sleep EEG slowing ratio over all derivations was mainly associated with lower FEOBV uptake in the medial temporal lobe (including the hippocampus and parahippocampal gyrus), amygdala, basal forebrain, insula, orbitofrontal and temporal cortex **(Fig. 3 and Supplementary Table 4)**. In men, higher REM sleep EEG slowing ratio over all derivations was associated with lower FEOBV uptake mainly in the frontal, temporal and parietal neocortex **(Fig. 3 and Supplementary Table 4)**.

**Figure 3.**
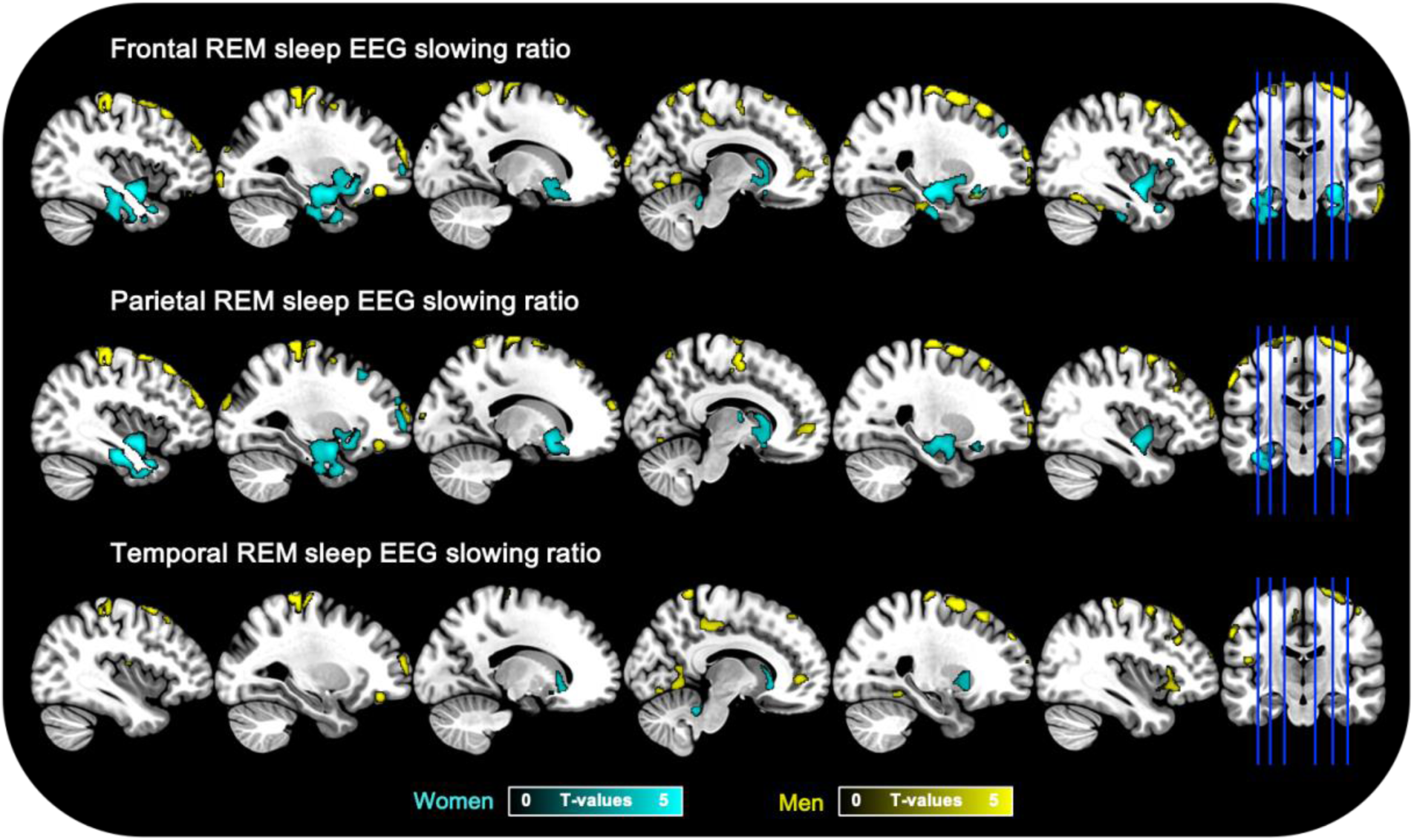
Differential associations between FEOBV uptake and REM sleep EEG slowing ratios in men and women. Negative voxel-wise regressions were performed between REM sleep EEG slowing ratio on F3-F4 (frontal, *n=22*), P3-P4 (parietal, *n=23*) and T3-T4 (temporal, *n=23*) derivations and FEOBV SUVR maps corrected for partial volume effects, controlling for age. Results are presented at the *P<0.005* uncorrected level, combined with a minimum cluster size of *k=100*. Detailed clusters statistics and peak coordinates are available in Supplementary Table 4.

## 4. Discussion

In this study, we provide the first direct *in vivo* evidence that brain cholinergic denervation is associated with REM sleep EEG slowing in older adults, even before the onset of dementia. This finding substantiates a long-standing hypothesis, suggesting that reduced cholinergic input to the cortex compromises the characteristic of EEG activity during REM sleep. This association was specific to REM sleep, as no significant relationship was found with NREM sleep or resting-state wakefulness EEG slowing, underscoring the cholinergic dependency of REM sleep. Preliminary analyses also revealed sex-specific effects in the relationship between reduced cholinergic innervation and REM sleep EEG slowing, which should be further clarified in future studies.

The fact that REM sleep is sensitive to cholinergic denervation aligns with prior research emphasizing the key role of the cholinergic system in REM sleep physiology. Both wakefulness and REM sleep are characterized by an intense cortical activation with desynchronized EEG patterns, characterized by low-amplitude fast-frequency EEG rhythms. However, unlike wakefulness where cortical activation arises from the combined influence of multiple neurotransmitter systems (e.g., cholinergic, noradrenergic, serotoninergic, orexinergic)^41,42^, REM sleep critically depends on cholinergic inputs, as other neurotransmitter systems are silenced during this sleep stage^22,23,41,42^. This cholinergic dependency is further supported by converging animal studies showing that pharmacological, optogenetic and lesion-induced experimental disruption of cholinergic neurons reduces REM sleep and selectively promotes EEG delta activity^43–47^. In addition, a double-blind randomized clinical trial has reported that increasing brain acetylcholine levels through the administration of anticholinesterase inhibitors such as donepezil reduce REM sleep EEG slowing in Alzheimer’s disease patients^48^. Our results extend these prior observations by demonstrating, for the first time, that in humans, reduced cortical cholinergic innervation is directly associated with REM sleep EEG slowing, even before dementia onset. Importantly, this association was specific to REM sleep, as no significant association was observed with EEG slowing computed during NREM sleep or resting-state wakefulness, even at a more permissive statistical threshold. While replication in a larger sample of participants is necessary, our findings support the idea that REM sleep quantitative EEG may constitute an early and specific marker of cholinergic denervation in older populations.

Cholinergic denervation likely results from the early accumulation of Alzheimer’s disease - related pathology, as cholinergic neurons are among the most vulnerable to the accumulation of abnormal proteins. This selective vulnerability may stem from the structural and functional properties of cholinergic neurons, as they exhibit large and long-distance projecting axons, complex branching and high metabolic rates^49,50^. Consistently, numerous post-mortem studies have reported that brainstem and basal forebrain cholinergic nuclei are early affected by intracellular tau pathology^1,51,52^. In addition to neurofibrillary tangles, cholinergic neurons have also been shown to be sensitive to amyloid toxicity^53–57^. *In vivo* structural neuroimaging studies confirm that Alzheimer’s disease patients present with a significant atrophy of basal forebrain nuclei, especially the Nucleus Basalis of Meynert^58–60^. Several other studies suggest that basal forebrain atrophy may also be detectable before the onset of dementia, in patients with Mild Cognitive Impairment (especially those who will convert to Alzheimer’s disease) and even subjective cognitive decline^59,61–63^. Longitudinal studies have shown that the atrophy of the basal forebrain may precede and predict the atrophy of the entorhinal cortex, also affected early by Alzheimer’s disease pathology^2,4^. Beyond the structural alteration of the basal forebrain, post-mortem studies show that several markers of cholinergic function are altered in the brain of Alzheimer’s disease patients, including a decline of acetylcholine levels, choline acetyltransferase and acetylcholinesterase activity^64–66^. Recent studies using FEOBV-PET have shown that patients with Alzheimer’s disease exhibit significant cholinergic denervation mainly in the temporo-parietal, medial prefrontal and cingulate cortices^13,14^, which correlate with the atrophy of the nucleus basalis of Meynert^3,14^. These results are in accordance with those of a recent study showing similar associations between cholinergic denervation and basal forebrain atrophy in Mild Cognitive Impairment patients^15^. Such cholinergic denervation may have important clinical consequences. For example, basal forebrain atrophy has been associated to cognitive decline and the severity of memory and attentional deficits in Alzheimer’s disease and Mild Cognitive Impairment patients^62,67,68^, and seems to predict conversion from Mild Cognitive Impairment to Alzheimer’s disease dementia^2,69^. Our findings extend this literature by demonstrating that cholinergic dysfunction not only affects cognition, but also relate to REM sleep alterations frequently observed in prodromal Alzheimer’s disease. This is particularly relevant given the essential role of REM sleep in memory processing, synaptic plasticity and emotion regulation^70,71^. Thus, in cognitively unimpaired older adults at increased risk for Alzheimer’s disease, or prodromal stages of the disease, cholinergic degeneration may impact cognition and psycho-affective health through both direct and sleep-mediated mechanisms.

Interestingly, our results provide preliminary insights into potential sex differences in the relationship between cholinergic denervation and REM sleep EEG slowing. In women, cholinergic innervation (mainly in the hippocampus, parahippocampal gyrus and amygdala) was higher than in men, and higher REM sleep EEG slowing was mainly associated with cholinergic denervation in the hippocampus, parahippocampal region, amygdala, basal forebrain, insula, orbitofrontal and temporal cortex. In men, REM sleep EEG slowing was rather associated with cholinergic denervation in the fronto-temporo-parietal neocortex. Although these results should be interpreted with caution given the small sample size, they echo emerging evidence suggesting sex-specific patterns of vulnerability to Alzheimer’s disease pathology^72,73^. Higher cholinergic innervation in women could reflect a compensatory upregulation of cholinergic innervation triggered by the early accumulation of Alzheimer’s disease pathology in cholinergic nuclei^74,75^. Supporting this interpretation, evidence for an up-regulation of the activity of the cholinergic system in hippocampal regions has been suggested previously in individuals with Mild Cognitive Impairment, to compensate for an early neuronal loss in other medial temporal lobe subregions, such as the entorhinal cortex^74,75^. The fact that this upregulation may be particularly evident in women could be due to higher medial temporal tau burden in the early stages of Alzheimer’s disease compared to men^72^.

The main strengths of the present study include the *in vivo* assessment of cholinergic denervation using FEOBV-PET, combined with quantitative REM sleep EEG analyses, in a well-characterized sample of older adults without dementia and moderate-to-severe obstructive sleep apnea. However, some limitations must be acknowledged, including the small sample size, the lack of Alzheimer’s disease biomarkers and the cross-sectional study design. However, given the complexity of protocols including an FEOBV-PET procedure, our sample size is in line with prior investigations in older populations^13–15,76,77^. Future studies should replicate these observations in a larger sample of participants with well-characterized Alzheimer’s disease biomarkers (i.e., amyloid and tau pathology levels), further clarify potential sex-specific effects, and assess whether REM sleep EEG slowing predicts the conversion to dementia using a longitudinal design.

In conclusion, our findings provide the first direct evidence that REM sleep EEG slowing is a sensitive and specific signature of cholinergic denervation in older adults. This work uncovers a key neurophysiological consequence of cholinergic degeneration, reinforcing the pivotal role of the cholinergic system in sustaining REM sleep cortical activity in humans. It highlights the importance of considering REM sleep alterations not only as a symptom of dementia, but as a possible window into early pathological changes. Clinically, quantitative REM sleep EEG may serve as a promising non-invasive marker for the early detection of neurodegenerative processes and a critical target for disease-modifying interventions in aging.

## 5. Data availability statement

Data used in the present study will be available upon request to the corresponding author.

## 6. Acknowledgements

The authors would like to thank all the participants of the study, as well as Sonia Frenette, Hélène Blais, Stéphane Frenette, Antonys Melek, Samantha Mombelli, Marie-Ève Martineau-Dussault, Etienne Aumont and Olga Fliaguine for their help.

