## Supplementary Material for "REM sleep EEG slowing reflects brain cholinergic denervation in aging and Mild Cognitive Impairment"

**Supplementary Table 1. Clusters showing significantly higher cholinergic uptake in women compared to men.**

**Supplementary Figure 1. Associations between REM sleep characteristics and demographic variables.**

**Supplementary Table 2. Clusters significantly associated with REM sleep EEG slowing ratios.**

**Supplementary Figure 2. Fully adjusted associations between REM sleep EEG slowing ratio and cholinergic uptake.**

**Supplementary Table 3. Associations between the frontal resting-state wakefulness EEG slowing ratio and FEOBV-SUVr maps.**

**Supplementary Table 4. Clusters significantly associated with REM sleep EEG slowing ratios in sex-stratified analyses.**

**<u>Supplementary Table 1.</u> Clusters showing significantly higher cholinergic uptake in women compared to men.**

| Cluster label | k | T | Cluster-level $p_{FWE\text{-}corr}$ | MNI coordinates x | y | z |
|---|---|---|---|---|---|---|
| **Unadjusted Two-sample analysis** | | | | | | |
| L hippocampus, middle temporal gyrus, superior temporal gyrus, supramarginal gyrus, Heschl gyrus, rolandic operculum, insula, superior temporal pole, putamen, basal forebrain, fusiform gyrus, lingual gyrus, parahippocampal gyrus, amygdala, thalamus, calcarine cortex, posterior cingulate cortex, precuneus | 7603 | 5.31 | **<0.001** | -32 | -18 | -12 |
| R hippocampus, parahippocampal gyrus, amygdala | 1691 | 4.99 | 0.18 | 28 | -33 | -3 |
| R lingual gyrus, calcarine cortex, superior occipital gyrus | 2577 | 4.86 | **0.048** | 14 | -92 | -8 |
| L middle temporal gyrus, inferior temporal gyrus, angular gyrus | 2768 | 4.28 | **0.036** | -45 | -74 | 20 |
| R supramarginal gyrus, inferior parietal gyrus, angular gyrus | 720 | 4.27 | 0.74 | 45 | -38 | 39 |
| R middle cingulate gyrus | 807 | 4.25 | 0.67 | 18 | -30 | 40 |
| L middle occipital gyrus | 477 | 4.25 | 0.91 | -27 | -82 | 8 |
| R insula, putamen, superior temporal gyrus, middle temporal gyrus, inferior temporal gyrus, supramarginal gyrus, Rolandic operculum, postcentral gyrus, precentral gyrus | 6777 | 4.24 | **<0.001** | 38 | -16 | 0 |
| L middle occipital gyrus, lingual gyrus, calcarine gyrus | 1316 | 4.14 | 0.33 | -10 | -94 | 0 |
| L middle temporal gyrus, superior temporal gyrus, middle temporal gyrus, temporal pole | 1100 | 4.09 | 0.45 | -48 | -14 | -16 |
| L inferior frontal gyrus, middle frontal gyrus | 1376 | 4.04 | 0.30 | -52 | 24 | 8 |
| R middle temporal gyrus | 463 | 4.03 | 0.92 | 52 | -66 | -3 |
| R lingual gyrus, precuneus, calcarine cortex, cuneus. | 656 | 3.95 | 0.79 | 20 | -54 | 0 |
| B basal forebrain, olfactory cortex, gyrus rectus, superior frontal gyrus | 1124 | 3.86 | 0.44 | 2 | 9 | -10 |
| L inferior parietal gyrus | 633 | 3.82 | 0.81 | -33 | -50 | 44 |
| L precentral gyrus | 181 | 3.66 | 1.00 | -21 | -26 | 54 |
| L postcentral gyrus | 422 | 3.64 | 0.94 | -44 | -16 | 33 |
| R inferior frontal gyrus, middle frontal gyrus | 405 | 3.62 | 0.95 | 56 | 24 | 15 |
| L inferior frontal gyrus | 192 | 3.60 | 1.00 | -40 | 9 | 26 |
| B superior frontal gyrus, L anterior cingulate cortex | 725 | 3.53 | 0.74 | -14 | 62 | -9 |
| R lingual gyrus | 137 | 3.46 | 1.00 | 21 | -58 | -4 |
| R inferior frontal gyrus | 107 | 3.42 | 1.00 | 44 | 12 | 27 |
| R postcentral gyrus | 116 | 3.35 | 1.00 | 32 | -39 | 58 |
| L middle frontal gyrus, L inferior frontal gyrus | 209 | 3.27 | 1.00 | -46 | 32 | 27 |
| L inferior temporal gyrus | 111 | 3.16 | 1.00 | -57 | -18 | -28 |
| **Age-adjusted Two-sample analysis** | | | | | | |
| L hippocampus, parahippocampal gyrus, lingual gyrus, amygdala | 1627 | 4.63 | 0.20 | -32 | -18 | -14 |
| L insula, Heschl gyrus, Rolandic operculum | 766 | 4.52 | 0.70 | -38 | -15 | 15 |
| R hippocampus, parahippocampal gyrus | 856 | 4.43 | 0.63 | 32 | -30 | -10 |
| R lingual gyrus, inferior occipital gyrus | 368 | 4.12 | 0.96 | 14 | -92 | -8 |
| B basal forebrain, olfactory gyrus | 369 | 3.88 | 0.96 | 2 | 8 | -9 |

| | | | | | | |
|---|---|---|---|---|---|---|
| R superior temporal gyrus, insula, Rolandic operculum. | 910 | 3.73 | 0.59 | 57 | -6 | 2 |
| R supramarginal gyrus | 149 | 3.66 | 1.00 | 58 | -27 | 32 |
| R superior temporal gyrus, middle temporal gyrus | 309 | 3.58 | 0.98 | 57 | -12 | -4 |
| L middle temporal gyrus | 467 | 3.48 | 0.92 | -56 | -56 | 8 |
| L superior temporal gyrus, Rolandic operculum, supramarginal gyrus | 506 | 3.45 | 0.90 | -57 | -34 | 22 |
| L superior temporal gyrus | 241 | 3.42 | 0.99 | -48 | 2 | -12 |
| L middle occipital gyrus, calcarine cortex | 101 | 3.39 | 1.00 | -10 | -94 | 0 |
| R superior temporal pole, amygdala, hippocampus | 138 | 3.32 | 1.00 | 28 | 3 | -22 |
| L inferior frontal gyrus, middle frontal gyrus, insula | 370 | 3.29 | 0.96 | -44 | 42 | -3 |

Results of two-sample voxel-wise comparisons showing higher cholinergic uptake in women *(n=14)* compared to men *(n=10)*, at the $P < 0.005$ uncorrected level combined with a minimum cluster size of $k=100$. Clusters surviving a cluster-level FWE correction are highlighted in bold. For each cluster, upper coordinates correspond to the statistical peak of the cluster. Abbreviations: FWE, family wise error; L, left; MNI, Montreal Neurological Institute; Nb, number; R, right.

**<u>Supplementary Figure 1.</u> Associations between REM sleep characteristics and demographic variables.**

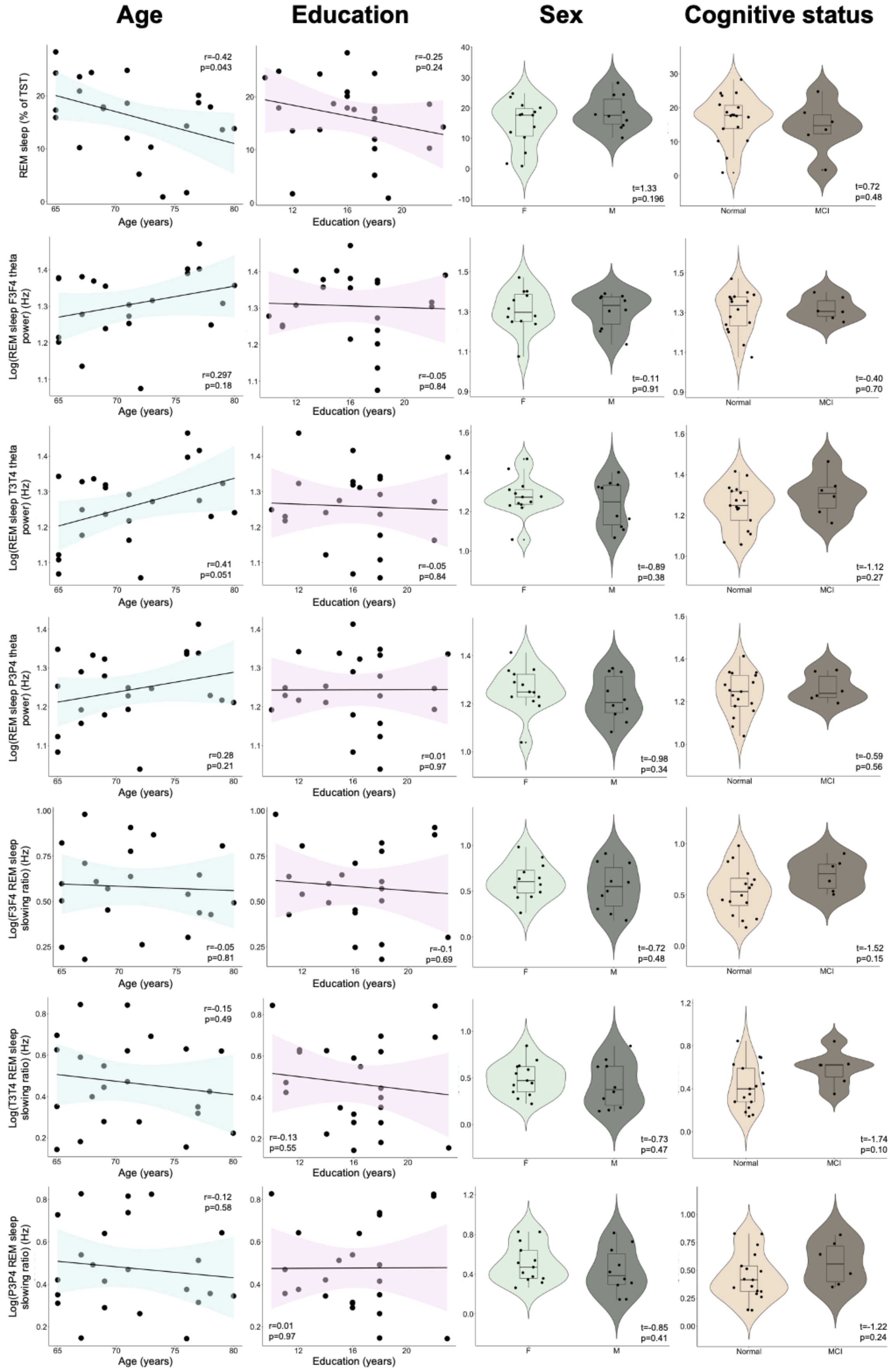

Associations between REM sleep percentage, theta power and EEG slowing ratios and demographic variables. Between-group differences in REM sleep outcomes according to sex or cognitive status were assessed using Student T-tests, and associations with age and education are assessed using Pearson's correlations. No association survived a false discovery rate correction for multiple comparisons. Abbreviations: Hz, Hertz; REM, rapid eye movement; TST, total sleep time.

**<u>Supplementary Table 2.</u> Clusters significantly associated with REM sleep EEG slowing ratios.**

| Cluster label | k | T-value | $p_{FWE\text{-}corrected}$ | MNI coordinates | | |
|---|---|---|---|---|---|---|
| | | | | x | y | z |
| **Frontal REM sleep EEG slowing ratio (n=22)** | | | | | | |
| B vermis, R lingual gyrus, B fusiform gyrus, B middle temporal gyrus, B superior temporal gyrus, B inferior temporal gyrus, B temporal pole, R parahippocampal gyrus, B hippocampus, B amygdala, B insula, R postcentral gyrus, R precentral gyrus, R supplementary motor area, R superior frontal gyrus, B middle frontal gyrus, R inferior frontal gyrus pars orbitalis, B superior frontal gyrus-medial orbital, R inferior frontal gyrus-opercular part, R inferior frontal gyrus-triangular part, B anterior cingulate cortex, B rectus gyrus, B medial orbital gyrus, B posterior orbital gyrus, L anterior orbital gyrus. | 38622 | 5.58 | <0.001 | 6 | -58 | -6 |
| B cuneus, L precuneus, L angular gyrus, L middle and superior occipital gyrus, L calcarine cortex. | 2667 | 5.51 | 0.014 | -6 | -78 | 28 |
| B precuneus, R superior parietal gyrus, L inferior parietal gyrus, B middle cingulate gyrus, B postcentral gyrus, L precentral gyrus, B paracentral lobule, B supplementary motor area, L superior frontal gyrus, L middle frontal gyrus. | 8201 | 4.84 | <0.001 | -14 | -46 | 72 |
| L superior frontal gyrus, L middle frontal gyrus, L inferior frontal gyrus-triangular part. | 5451 | 4.48 | <0.001 | -2 | 45 | 45 |
| **Parietal REM sleep EEG slowing ratio (n=23)** | | | | | | |
| B cuneus, B precuneus, B superior occipital gyrus, L middle occipital gyrus, B fusiform gyrus, B postcentral gyrus, B superior parietal gyrus, L inferior parietal gyrus, B precentral, B superior frontal gyrus, B middle frontal gyrus, B inferior frontal gyrus-opercular part, B inferior frontal gyrus-triangular part, B inferior frontal gyrus pars orbitalis, R Rolandic operculum, B supplementary motor area, B olfactory cortex, B superior frontal gyrus-medial and medial orbital parts, B rectus gyrus, B orbital gyrus (anterior, medial and posterior parts), R anterior cingulate cortex, B insula, B middle cingulate gyrus, L hippocampus, L amygdala, B paracentral lobule, L putamen, R Heschls gyrus, B superior temporal gyrus, B temporal pole, B middle temporal gyrus, B inferior temporal gyrus, B cerebellar hemispheres. | 51051 | 4.25 | <0.001 | -6 | -78 | 28 |
| B vermis, R fusiform gyrus, R lingual gyrus, R parahippocampal gyrus. | 2170 | 3.72 | 0.039 | 4 | -60 | -6 |

Whole-brain voxel-wise negative multiple regression analyses were performed between REM sleep EEG slowing ratios on frontal (averaged F3-F4) and parietal (averaged P3-P4) derivations, and FEOBV SUVR maps. Results are presented at the $P < 0.005$ (uncorrected) level combined with a FWE cluster-level correction, controlling for sex. For each cluster, upper coordinates correspond to the statistical peak of the cluster. <u>Abbreviations:</u> B, bilateral; FWE, family wise error; L, left; MNI, Montreal Neurological Institute; R, right; REM, rapid eye movement.

**<u>Supplementary Figure 2.</u> Fully adjusted associations between REM sleep EEG slowing ratio and cholinergic uptake.**

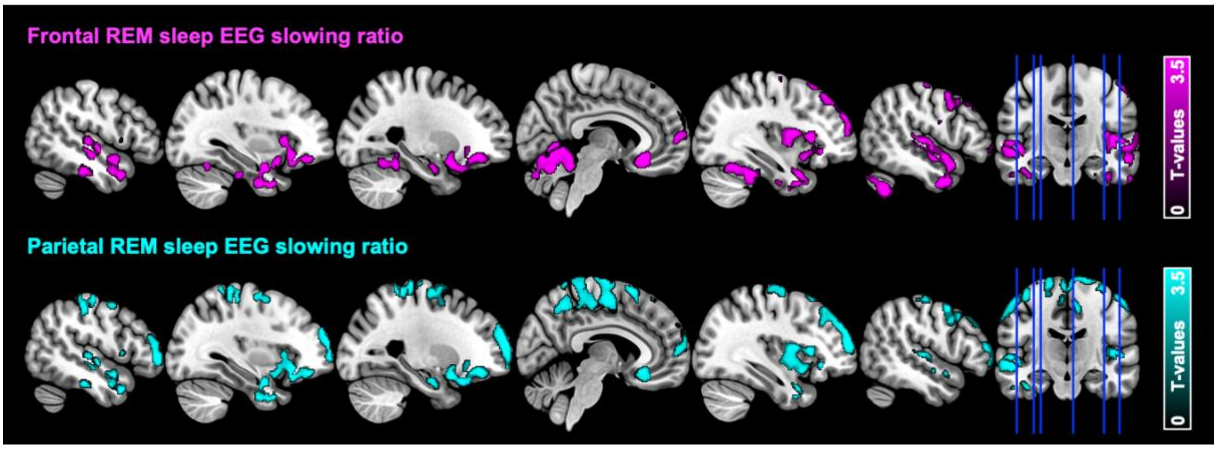


Whole-brain negative voxel-wise regressions between REM sleep EEG slowing ratio on F3-F4 (top, *n=22*) and P3-P4 (bottom, *n=23*) derivations and FEOBV-SUVR maps corrected for partial volume effects, controlling for age, sex and cognitive status. Results are presented at the $P < 0.005$ uncorrected level, combined with a cluster-level FWE correction. <u>Abbreviations</u>: FEOBV, [$^{18}$F]-Fluoroethoxybenzovesamicol; FWE, family wise error; SUVR, standard uptake value ratio, REM, rapid eye movement.

**Supplementary Table 3. Association between the frontal resting-state wakefulness EEG slowing ratio and FEOBV-SUVR maps.**

| Cluster label | k | T-value | $p_{FWE\text{-}corrected}$ | MNI coordinates | | |
|---|---|---|---|---|---|---|
| | | | | x | y | z |
| Right superior temporal pole | 107 | 4.38 | 1.000 | 36 | 9 | -27 |
| Left superior frontal gyrus | 193 | 3.98 | 0.997 | -26 | 15 | 51 |
| Left superior frontal gyrus | 137 | 3.79 | 0.999 | -26 | 39 | 44 |

Whole-brain voxel-wise negative multiple regression between the resting-state wakefulness EEG slowing ratio computed on frontal derivations (averaged F3-F4, $n=18$) and FEOBV-SUVR maps, controlling for sex. Results are presented at the $P < 0.005$ (uncorrected) level, combined with a minimum cluster size of $k=100$. Abbreviations: FEOBV, [$^{18}$F]-Fluoroethoxybenzovesamicol; FWE, family wise error; SUVR, standard uptake value ratio; REM, rapid eye movement.

**<u>Supplementary Table 4.</u> Clusters significantly associated with REM sleep EEG slowing ratios in sex-stratified analyses.**

| Group | Cluster label | k | T-value | $p_{FWE\text{-}corrected}$ | MNI coordinates | | |
|---|---|---|---|---|---|---|---|
| | | | | | x | y | z |
| **Frontal REM sleep EEG slowing ratio (n=22)** | | | | | | | |
| Women | Cerebellar vermis | 220 | 7.80 | 0.995 | 6 | -40 | -21 |
| | **R hippocampus, parahippocampal gyrus, putamen, amygdala, basal forebrain, nucleus accumbens, gyrus rectus, medial orbital gyrus** | **3651** | **1.36** | **0.003** | **27** | **-20** | **-15** |
| | **L insula, superior temporal gyrus, inferior temporal gyrus, middle temporal gyrus, amygdala, hippocampus, putamen, fusiform gyrus, basal forebrain, gyrus rectus, medial orbital gyrus** | **5307** | **6.88** | **<0.001** | **-39** | **-9** | **-15** |
| | L anterior cingulate cortex | 142 | 4.55 | 0.999 | -2 | 38 | 6 |
| | L superior frontal gyrus | 133 | 4.41 | 1.000 | -24 | 58 | 4 |
| | R superior frontal gyrus | 101 | 4.16 | 1.000 | 22 | 44 | 38 |
| Men | B lingual gyrus | 582 | 8.98 | 0.296 | 6 | -62 | -2 |
| | L precentral gyrus, supplementary motor area, precuneus, paracentral lobule, superior parietal gyrus, postcentral gyrus | 458 | 8.80 | 0.511 | -22 | -22 | 72 |
| | L postcentral gyrus, precentral gyrus, paracentral lobule | 624 | 8.27 | 0.243 | -33 | -30 | 58 |
| | **R superior frontal gyrus, precentral gyrus** | **2140** | **7.84** | **<0.001** | **27** | **27** | **56** |
| | R middle frontal gyrus | 322 | 7.12 | 0.816 | 42 | 8 | 56 |
| | L superior frontal gyrus | 293 | 7.06 | 0.873 | -14 | 38 | 54 |
| | R precentral gyrus, postcentral gyrus | 180 | 6.83 | 0.992 | 63 | -2 | 27 |
| | R superior parietal gyrus, precuneus, paracentral lobule | 818 | 6.74 | 0.097 | 15 | -63 | 68 |
| | L postcentral gyrus, supramarginal gyrus | 294 | 6.63 | 0.871 | -57 | -14 | 46 |
| | L supplementary motor area, paracentral lobule, middle cingulate cortex | 182 | 6.53 | 0.991 | -10 | -22 | 50 |
| | R superior frontal gyrus | 169 | 6.23 | 0.995 | 27 | 62 | 15 |
| | L orbitofrontal cortex | 221 | 6.02 | 0.969 | -27 | 39 | -14 |
| | L middle frontal gyrus | 242 | 5.81 | 0.949 | -50 | 21 | 38 |
| | L middle temporal gyrus, angular gyrus, superior temporal gyrus | 754 | 5.71 | 0.132 | -64 | -32 | 3 |
| | R superior frontal gyrus | 233 | 5.68 | 0.958 | 20 | 66 | -8 |
| | R superior frontal gyrus | 191 | 5.62 | 0.988 | 8 | 56 | -2 |
| | L precuneus, superior parietal gyrus | 243 | 5.59 | 0.948 | -33 | 24 | 51 |
| | L precuneus, superior parietal gyrus, paracentral lobule | 447 | 5.59 | 0.534 | -4 | -42 | 74 |
| | L supplementary motor area, L superior frontal gyrus | 113 | 5.56 | 1.000 | -12 | 22 | 62 |
| | R supplementary motor area | 102 | 5.45 | 1.000 | 6 | 18 | 66 |
| | R fusiform gyrus | 237 | 5.43 | 0.954 | 30 | -33 | -26 |
| | L middle occipital gyrus, inferior occipital gyrus | 152 | 5.42 | 0.998 | -28 | -94 | -2 |
| | L precentral gyrus, middle frontal gyrus, postcentral gyrus | 359 | 5.30 | 0.734 | -42 | -6 | 62 |
| | R supplementary motor area | 123 | 5.26 | 1.000 | 8 | -6 | 57 |
| | L orbitofrontal cortex | 110 | 5.20 | 1.000 | -22 | 15 | -22 |
| | L superior frontal gyrus, middle frontal gyrus | 586 | 5.17 | 0.290 | -28 | 56 | 18 |
| | R fusiform gyrus | 116 | 5.10 | 1.000 | 39 | -57 | -20 |
| | R cuneus | 159 | 5.07 | 0.997 | 14 | -102 | 9 |
| | R orbitofrontal cortex | 104 | 4.91 | 1.000 | 26 | 20 | -18 |

| | | | | | | | |
|---|---|---|---|---|---|---|---|
| | R middle temporal gyrus, inferior temporal gyrus, superior temporal gyrus | 715 | 4.87 | 0.158 | 66 | -20 | -21 |
| | L rectus gyrus | 117 | 4.77 | 1.000 | -6 | 26 | -18 |
| | R superior temporal gyrus | 104 | 4.72 | 1.000 | 64 | -33 | 4 |
| | L superior occipital gyrus | 211 | 4.66 | 0.997 | -21 | -86 | 36 |
| | L inferior temporal gyrus | 100 | 4.64 | 1.000 | -50 | -60 | -16 |
| | R superior occipital gyrus, cuneus | 159 | 4.49 | 0.997 | 22 | -88 | 28 |
| | B cuneus | 102 | 4.39 | 1.000 | 4 | -84 | 26 |
| | L inferior temporal gyrus | 134 | 4.29 | 0.999 | -45 | -42 | -27 |
| | R lingual gyrus, fusiform gyrus | 103 | 4.17 | 1.000 | 20 | -50 | -9 |
| **Parietal REM sleep EEG slowing ratio (n=23)** | | | | | | | |
| Women | **L inferior temporal gyrus, insula, hippocampus, amygdala, parahippocampal gyrus, fusiform, inferior temporal gyrus, temporal pole** | **5426** | **6.42** | **<0.001** | **-39** | **-20** | **-22** |
| | Cerebellar vermis | 135 | 5.17 | 0.999 | 3 | -40 | -20 |
| | R insula, putamen, hippocampus, amygdala, parahippocampal gyrus | 1719 | 4.90 | 0.097 | 36 | -3 | -10 |
| | R thalamus, nucleus accumbens, basal forebrain, caudate nucleus, gyrus rectus, medial orbital gyrus | 703 | 4.23 | 0.664 | 12 | -4 | 12 |
| | L superior frontal gyrus | 319 | 4.21 | 0.972 | -26 | 58 | 4 |
| | L middle frontal gyrus | 103 | 3.87 | 1.000 | -26 | 24 | 45 |
| Men | L precentral gyrus, postcentral gyrus | 850 | 10.45 | 0.136 | -34 | -30 | 60 |
| | R superior frontal gyrus, precentral gyrus, supplementary motor area | 816 | 8.90 | 0.156 | 20 | -15 | 75 |
| | L postcentral gyrus, supramarginal gyrus | 276 | 3.83 | 0.936 | -58 | -15 | 45 |
| | L precentral gyrus, paracentral lobule, supplementary motor area | 407 | 6.63 | 0.718 | -21 | -22 | 74 |
| | R superior frontal gyrus, middle frontal gyrus | 395 | 6.37 | 0.742 | 26 | 63 | 15 |
| | L superior frontal gyrus | 187 | 3.55 | 0.993 | -8 | 38 | 57 |
| | L superior frontal gyrus | 108 | 3.54 | 1.000 | 27 | 28 | 56 |
| | R supplementary motor area | 181 | 3.53 | 0.995 | 9 | -6 | 58 |
| | L precuneus, paracentral lobule, superior parietal gyrus | 417 | 6.18 | 0.698 | -4 | -40 | 72 |
| | L supplementary motor area, paracentral lobule, middle cingulate gyrus | 148 | 6.11 | 0.999 | -10 | -22 | 50 |
| | L middle frontal gyrus, inferior frontal gyrus | 398 | 6.09 | 0.736 | -34 | 26 | 51 |
| | L precentral gyrus, middle frontal gyrus | 296 | 5.98 | 0.912 | -50 | -2 | 52 |
| | L superior frontal | 130 | 5.92 | 1.000 | -21 | 2 | 70 |
| | R middle frontal gyrus, inferior frontal gyrus | 470 | 5.75 | 0.592 | 44 | 28 | 38 |
| | R superior parietal gyrus, precuneus | 120 | 5.65 | 1.000 | 15 | -63 | 68 |
| | R middle frontal gyrus | 183 | 5.59 | 0.994 | 38 | 6 | 60 |
| | L superior frontal gyrus | 322 | 5.32 | 0.874 | -22 | 63 | 9 |
| | B lingual gyrus, cerebellar vermis | 333 | 5.22 | 0.856 | 6 | -60 | -2 |
| | L middle temporal gyrus, superior temporal gyrus | 234 | 5.18 | 0.973 | -66 | -30 | 2 |
| | L superior frontal gyrus | 100 | 5.15 | 1.000 | -15 | 63 | 20 |
| | R superior frontal gyrus | 136 | 5.03 | 0.999 | 9 | 54 | -2 |
| | L superior occipital gyrus, middle occipital gyrus | 317 | 4.54 | 0.882 | -21 | -87 | 36 |
| | L middle frontal gyrus, orbitofrontal cortex | 114 | 4.50 | 1.000 | -27 | 38 | -12 |
| **Temporal REM sleep EEG slowing ratio (n=23)** | | | | | | | |
| Women | Cerebellar vermis | 287 | 7.26 | 0.985 | 8 | -44 | -26 |
| | L anterior cingulate cortex, nucleus accumbens, basal forebrain, putamen, caudate nucleus | 387 | 4.41 | 0.953 | -6 | 22 | -6 |
| | R caudate nucleus | 173 | 4.37 | 0.998 | 14 | 21 | -8 |

| | | | | | | | |
|---|---|---|---|---|---|---|---|
| | R superior temporal pole | 105 | 4.06 | 1.000 | 50 | 4 | -12 |
| | R putamen | 282 | 3.70 | 0.986 | 30 | 18 | 3 |
| Men | Cerebellar vermis, B lingual gyrus, R calcarine cortex | 595 | 7.82 | 0.387 | 4 | -63 | -3 |
| | L postcentral gyrus, supramarginal gyrus, precentral gyrus | 303 | 7.49 | 0.906 | -57 | -12 | 45 |
| | L postcentral gyrus, precentral gyrus | 451 | 6.83 | 0.639 | -28 | -34 | 68 |
| | L middle frontal gyrus, inferior frontal gyrus | 468 | 6.70 | 0.606 | -51 | 20 | 36 |
| | R middle frontal gyrus, superior frontal gyrus, inferior frontal gyrus | 650 | 6.22 | 0.313 | 42 | 28 | 33 |
| | R postcentral gyrus | 106 | 6.18 | 1.000 | 63 | -2 | 28 |
| | R superior frontal gyrus, middle frontal gyrus | 559 | 6.17 | 0.442 | 24 | 4 | 62 |
| | R precentral gyrus | 117 | 6.13 | 1.000 | 42 | -22 | 64 |
| | R superior frontal gyrus | 136 | 5.90 | 0.999 | 16 | 36 | 54 |
| | R superior frontal gyrus, middle frontal gyrus | 104 | 5.77 | 1.000 | 26 | 50 | 38 |
| | L precentral gyrus, paracentral lobule | 153 | 5.61 | 0.999 | -26 | -24 | 68 |
| | R inferior parietal gyrus | 142 | 5.55 | 0.999 | 48 | -45 | 56 |
| | L superior frontal gyrus | 137 | 5.48 | 0.999 | -27 | 58 | 12 |
| | R precuneus | 122 | 5.45 | 1.000 | 8 | -50 | 72 |
| | R precentral gyrus, superior frontal gyrus | 314 | 5.02 | 0.890 | 26 | -20 | 72 |
| | R inferior frontal gyrus, insula | 362 | 4.39 | 0.811 | 44 | 26 | 3 |

Whole-brain voxel-wise negative multiple regression analyses were performed between REM sleep EEG slowing ratios on frontal (averaged F3-F4), temporal (averaged T3-T4) and parietal (averaged P3-P4) derivations and FEOBV-SUVR maps in groups stratified by sex. Results are presented at the $P < 0.005$ (uncorrected) level combined with a minimum cluster size of $k=100$, controlling for age. Clusters surviving a cluster-level FWE correction are highlighted in bold. For each cluster, coordinates correspond to the statistical peak of the cluster. Abbreviations: B, bilateral; FWE, family wise error; L, left; MNI, Montreal Neurological Institute; R, right; REM, rapid eye movement.
